# The International Cardiac Arrest Research (I-CARE) Consortium Electroencephalography Database

**DOI:** 10.1101/2023.08.28.23294672

**Authors:** Edilberto Amorim, Wei-Long Zheng, Mohammad M. Ghassemi, Mahsa Aghaeeaval, Pravinkumar Kandhare, Vishnu Karukonda, Jong Woo Lee, Susan T. Herman, Adithya Sivaraju, Nicolas Gaspard, Jeannette Hofmeijer, Michel J. A. M. van Putten, Reza Sameni, Matthew A. Reyna, Gari D. Clifford, M. Brandon Westover

## Abstract

**Objective:** To develop a harmonized multicenter clinical and electroencephalography (EEG) database for acute hypoxic-ischemic brain injury research involving patients with cardiac arrest.

**Design:** Multicenter cohort, partly prospective and partly retrospective.

**Setting:** Seven academic or teaching hospitals from the U.S. and Europe.

**Patients:** Individuals aged 16 or older who were comatose after return of spontaneous circulation following a cardiac arrest who had continuous EEG monitoring were included.

**Interventions:** not applicable.

**Measurements and Main Results:** Clinical and EEG data were harmonized and stored in a common Waveform Database (WFDB)-compatible format. Automated spike frequency, background continuity, and artifact detection on EEG were calculated with 10 second resolution and summarized hourly. Neurological outcome was determined at 3-6 months using the best Cerebral Performance Category (CPC) scale. This database includes clinical and 56,676 hours (3.9 TB) of continuous EEG data for 1,020 patients. Most patients died (N=603, 59%), 48 (5%) had severe neurological disability (CPC 3 or 4), and 369 (36%) had good functional recovery (CPC 1-2). There is significant variability in mean EEG recording duration depending on the neurological outcome (range 53-102h for CPC 1 and CPC 4, respectively). Epileptiform activity averaging 1 Hz or more in frequency for at least one hour was seen in 258 (25%) patients (19% for CPC 1-2 and 29% for CPC 3-5). Burst suppression was observed for at least one hour in 207 (56%) and 635 (97%) patients with CPC 1-2 and CPC 3-5, respectively.

**Conclusions:** The International Cardiac Arrest Research (I-CARE) consortium database provides a comprehensive real-world clinical and EEG dataset for neurophysiology research of comatose patients after cardiac arrest. This dataset covers the spectrum of abnormal EEG patterns after cardiac arrest, including epileptiform patterns and those in the ictal-interictal continuum.

## Introduction

More than 6 million cardiac arrests happen every year worldwide, with survival rates ranging from 1% to 10% depending on geographic location(1, 2). Severe brain injury is the most common cause of death for patients surviving initial resuscitation, with most survivors admitted to an intensive care unit (ICU) being comatose (3). Clinicians are typically asked during the first few days following cardiac arrest to monitor for seizures and offer a prognosis, i.e., determine the probability that the patient will have a poor outcome or eventually recover neurological function. Many abnormal EEG patterns seen in these patients do not meet criteria for electrographic seizures, but they may lay in the ictal interictal continuum and may require specific management guided by the EEG data (4). A poor prognosis prediction typically leads to the withdrawal of life sustaining therapies and death (5). However, reports of neurological recovery despite an initial poor prognosis exist in literature, underscoring the need of multimodal and thoughtful evaluation of these patients (6–8).

Clinical dilemmas about seizure management and prognostication emphasize the role of brain monitoring with electroencephalography (EEG) to provide direct assessment of brain function post-cardiac arrest (7, 9). Specific EEG signatures reflect brain injury or potential for neurological recovery. Clinical neurophysiologists have come to recognize numerous patterns of brain activity that help predict outcomes following cardiac arrest, including the presences of burst suppression (alternating periods of high and low voltage), seizures, or status epilepticus. Several studies have shown that the longitudinal evolution of these EEG patterns and transition between them may provide additional prognostic information (10–14). However, qualitative interpretation of continuous EEG is laborious, expensive, and requires review from neurologists with advanced training in neurophysiology who are unavailable in most medical centers.

Automated analysis of continuous EEG data has the potential to improve prognostic accuracy and to increase access to brain monitoring where experts are not readily available (12, 13, 15–18). However, the datasets used in most studies typically involve small numbers of patients (<100) from single medical centers, which are unsuitable for high-quality machine learning analyses (7). To overcome this limitation the International Cardiac Arrest REsearch consortium (I-CARE) assembled a large representative set of clinical and EEG data from comatose patients with cardiac arrest who underwent continuous EEG monitoring following cardiac arrest and had neurologic outcomes assessed.

## Methods

This database consists of retrospectively obtained clinical and EEG data from four academic hospitals in the United States (Beth Israel Deaconess Medical Center, Brigham and Women’s Hospital, Massachusetts General Hospital, and Yale New Haven Medical Center, and prospectively collected data from two hospitals in the Netherlands (Medisch Spectrum Twente and Rinjstate Hospital) and one from Belgium (Erasme Hospital). Data from the two hospitals from the Netherlands was analyzed as a single institution as both hospitals and investigators are affiliated with the same University. Independent Institutional Review Board approvals at participating hospitals were pursued. Need for informed consent was waived by the institutional review board at participating centers for analysis of data obtained as part of routine medical care.

## Local Data Deidentification

Each individual hospital pursued deidentification of the clinical and EEG data locally (Figure 1). To protect patient privacy, all ages above 89 were aggregated into a single category and encoded as “90”. Each institution shared the data to a central repository (bdsp.io).

**Figure 1:**
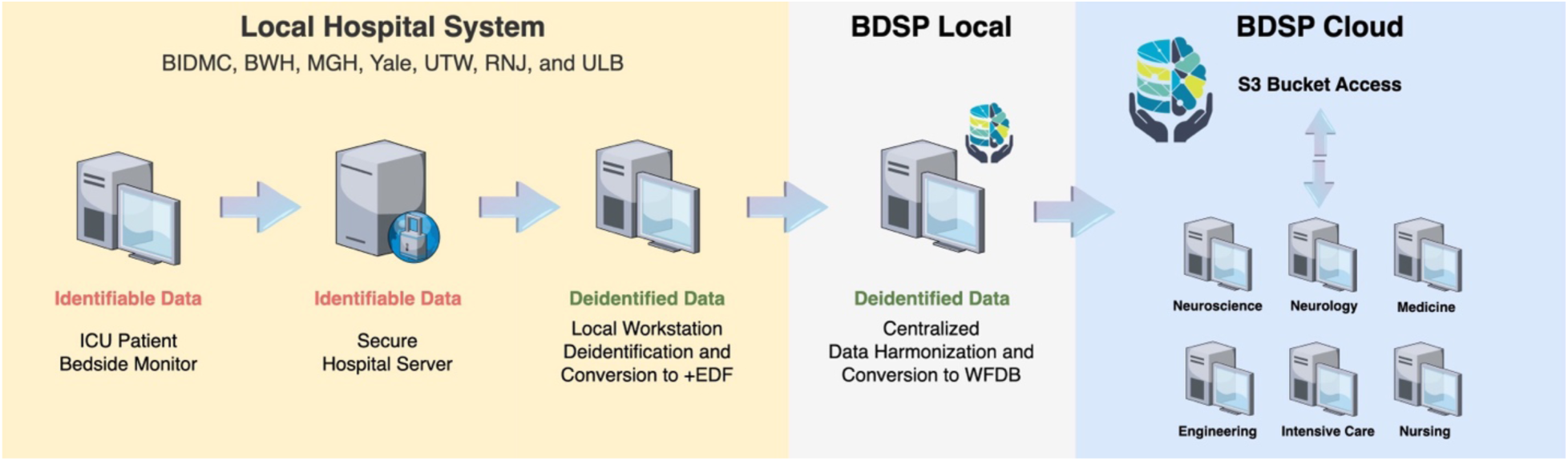
Clinical and EEG data processing and data sharing.

## Central Data Processing and Cloud Sharing

Data from individual hospitals were centrally harmonized. First, the .edf files from each center were converted using MATLAB software 2022 and EEGLAB Toolbox (v.2022.1) to .mat files. Then, the .mat EEG files and clinical data were converted into the Waveform Database (WFDB)-compatible format with 16-bit representation for the signal data. Sampling rate ranged from 200 Hz to 2,040 Hz. No filtering was performed in files included in the database. The EEG data was timed based on the time of ROSC. The final files were uploaded to a central cloud repository using an Amazon S3 object storage. This system allows for open data access and analysis using a secure and auditable cloud environment.

## Clinical Data

Patients 16 years or older with out-of-hospital or in-hospital cardiac arrest who had return of spontaneous circulation (ROSC). All comatose patients included were admitted to an ICU and underwent EEG monitoring (coma is defined as Glasgow Coma Scale <9 or inability to follow commands). The EEG monitoring is typically started within hours of cardiac arrest and continues for several hours to days depending on the patients’ condition, therefore recording start time and duration vary from patient to patient. Patient information includes information recorded at the time of admission (age, sex), location of arrest (out or in-hospital), and type of cardiac rhythm recorded at the time of resuscitation (shockable rhythms include ventricular fibrillation or ventricular tachycardia and non-shockable rhythms include asystole and pulseless electrical activity, or unknown). The time between cardiac arrest and ROSC is provided for patients who had that information recorded by the emergency medical services team. Patient temperature after cardiac arrest is controlled using a closed-loop feedback device (TTM) in most patients. The temperature level can be set at 33 degrees Celsius, 36 degrees Celsius, or no set temperature. Sedation and analgesia were used as needed by treating clinicians. Sedatives used and typical dose ranges were: propofol (25–80 mcg/kg/min), midazolam (0.1-0.7 mg/kg/h), or fentanyl (25– 200 mcg/h). Propofol was the initial sedative of choice for six hospitals and one institution used midazolam instead. Neuromuscular blockade during TTM initiation was only systematically used in one institution, with the remaining hospitals using neuromuscular blockade as needed.

## Patient Outcomes

Neurological outcome was determined prospectively in two hospitals from the Netherlands by phone interview (at 6 months from ROSC). For the remaining five hospitals, neurological outcome was retrospectively determined through chart review (at 3-6 months from ROSC). Neurological function was determined using the best Cerebral Performance Category (CPC) scale (19). In the five hospitals without prospective follow up, patients who achieved good neurological function (CPC score of 1 or 2) by the time of hospital discharge were considered to have achieved their best CPC score and no additional chart review was performed. The CPC is an ordinal scale ranging from 1 to 5:

CPC = 1: good neurological function and independent for activities of daily living.

CPC = 2: moderate neurological disability but independent for activities of daily living.

CPC = 3: severe neurological disability.

CPC = 4: unresponsive wakefulness syndrome (previously known as vegetative state).

CPC = 5: dead.

## EEG data

Each EEG file contains an array with EEG signals from 19 channel recorded using the 10-20 international system that has been harmonized. Additional channels included in the recordings (e.g., additional reference or EKG channels) may be present for some patients and they have been included as well. Patients may need to have brain monitoring interrupted transiently while in the ICU, so gaps in the data can be present. The EEG recordings can continue for hours to days, so the EEG signals are prone to quality deterioration from non-physiological artifacts. It is routine practice to continue EEG for 24-72h, but recordings may be abbreviated or prolonged if patients regain consciousness, have withdrawal of life-sustaining therapies, die, or develop seizures seizures.

## Quantitative EEG analysis

To illustrate EEG data trends across outcomes for participating hospitals included in this database, we acquired two quantitative EEG (QEEG) features representing epileptiform activity frequency (spike frequency) and background continuity (background continuity index [BCI]).(20) Prior to QEEG feature analysis, The EEG signals were resampled to 100 Hz and underwent frequency filtering (0.5-50 Hz). Independent component analysis was applied to attenuate artifacts prior to averaging features for consecutive 10-second epochs (EEGLAB Toolbox, v.2022.1). Artifact was detected for individual channels for every consecutive five seconds of EEG data. There were two types of EEG artifacts acquired: 1) overly high signal amplitude (average five-second amplitude above 500 µV or any signal with amplitude above 900 µV for more than 0.1 second) and 2) presence of a flat an invariant background (signal value with standard deviation < 0.2 µV for more than two seconds). Five-second epochs without any type of artifact detected were considered artifact-clean. For each hour, patients with less than 50% of the EEG data considered artifact-clean had the non-artifactual data averaged and used for analyses. Segments with 50% or more of artifactual data were excluded from the QEEG analysis.

The BCI reflects the fraction of EEG not spent in suppression and ranges between 0 and 1. Suppressions were defined as segments with amplitudes of <10 μV for at least 0.5 seconds. Spike frequency was determined based on the number of epileptiform discharges detected in consecutive five-minute epoch using a modified version of the automated spike detection algorithm (SpikeNet) (15). The SpikeNet algorithm has a 0.5 Hz resolution, therefore discrete wavelet transform followed by peak detection was pursued to identify additional spikes within a half second window (*findpeaks*) using the MATLAB signal processing toolbox. A dynamic amplitude threshold based on the signal distribution at each 500 ms window that had a spike detected using SpikeNet was applied to determine the spike frequency. We used the one-hour average QEEG value for artifact-clean data to classify the QEEG data in one of five categories based on: 1) the presence of spikes with 1 Hz or more frequency (category: epileptiform activity present) or 2) across four levels of background continuity (continuous [BCI >90%], discontinuous [BCI 50-90%], burst suppression [BCI 10-50%], and suppressed [BCI <10%]). Patients classified under the epileptiform activity category were not included in the four continuity categories, therefore each individual patient could only be included into a single category at each hour. Code for feature acquisition is available in an online repository (https://github.com/bdsp-core).

## Algorithms

The I-CARE dataset was used as part of the George B. Moody PhysioNet Challenge 2023 (21). The PhysioNet Challenges are annual data science competitions that invite international teams from academia and industry to develop algorithmic, open-source approaches for addressing unsolved problems. The PhysioNet Challenge 2023 asked teams to use the I-CARE dataset to predict the neurological recovery of patients at 72 hours after ROSC.

## Results

One thousand and twenty patients were included with a total of 56,676 hours of EEG data (Figure 2 and Figure S1). Six hundred and three patients died (59%), 48 (5%) survived with severe neurological disability (CPC 3 or 4), and 369 (36%) had good functional recovery (CPC 1-2). The median time to EEG initiation from the time of ROSC was 14 (IQR: 15) hours. Mean duration of EEG recording was longer for patients with CPC 3 or CPC 4 (71-102 hours) compared to patients with CPC 1, CPC 2, or CPC 5 (53-61 hours). Most patients (77%) underwent TTM with a goal of 33 degrees Celsius. Patient characteristics and EEG recording information are summarized in Table 1 and Table S1.

**Figure 2:**
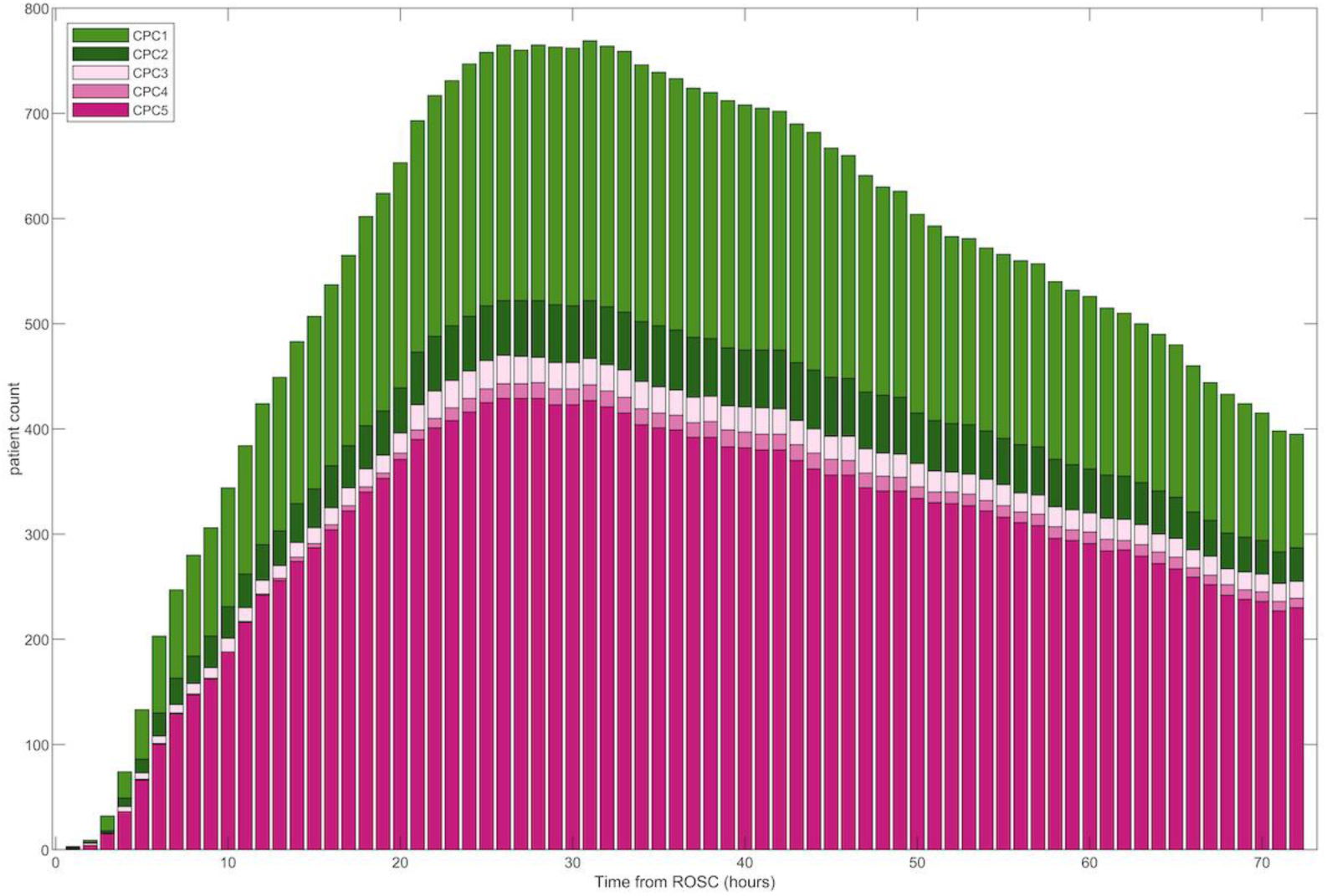
EEG data availability per hour from the time of ROSC stratified by neurologic outcome (CPC). CPC: Cerebral Performance Category; ROSC: return of spontaneous circulation.

**Table 1.** Patient Demographics Stratified by Neurological Outcome.

|  | **CPC 1** | **CPC 2** | **CPC 3** | **CPC 4** | **CPC5** |
| --- | --- | --- | --- | --- | --- |
| Number of subjects | 300 | 69 | 31 | 17 | 603 |
| Age (years, mean, median, std, iqr) | 57,58,15,20 | 56,57,14,17 | 66,69,11,16 | 53,53,20,37 | 62,65,16,21 |
| Female (%) | 88 (29) | 17 (25) | 11 (35) | 8 (47) | 198 (32) |
| Shockable Rhythm (%) | 214 (71) | 47 (68) | 13 (41) | 7 (41) | 188 (31) |
| TTM 33 (%) | 232 (77) | 55(79) | 21 (67) | 11 (64) | 468 (78) |
| TTM 36 | 28 (0.1) | 6 (0.09) | 5 (16) | 0 (0) | 41 (0.07) |
| No TTM | 40 (0.13) | 8 (0.12) | 5 (16) | 6 (35) | 94 (16) |
| EEG start time from ROSC (hours, mean, median, STD, IQR) | 15,13,12,14 | 13,12,10,15 | 15,14,13,14 | 19,20,6,8 | 17,14,13,14 |
| EEG duration (hours, mean, median, STD, IQR) | 53,47,35,35 | 61,48,41,27 | 71,65,51,39 | 102,110,60,98 | 54,46,40,45 |

CPC: Cerebral Performance Category
ROSC: return of spontaneous circulation
STD: standard deviation
IQR: interquantile range

Epileptiform activity was present for at least one hour in 258 patients (Figure 3). Time to start of epileptiform activity ranged from 4 to 72 hours from time of cardiac arrest. Most patients with epileptiform activity had a poor neurological outcome, but 69 (19%) of patients with CPC of 1-2 had epileptiform activity. In contrast, 177 (64%) patients with a continuous background within 24h had good neurological recovery. Burst suppression was observed for at least one hour in 150 (41%) and 378 (58%) of patients with CPC 1-2 and CPC 3-5 and a suppressed background in 57 (16%) and 257 (40%), respectively. One in four patients (N=96) with burst suppression beyond 24h from the time of ROSC had a good outcome. Prevalence of epileptiform activity and burst suppression or suppression varied among centers (Figure S1). The burden of artifactual data varied across time, going from 9.4% within 24h to 6.3%at 24-48h, and 4.7% after 48h. A summary of the incidence of EEG epileptiform activity and the BCI is shown in Table 2.

**Figure 3:**
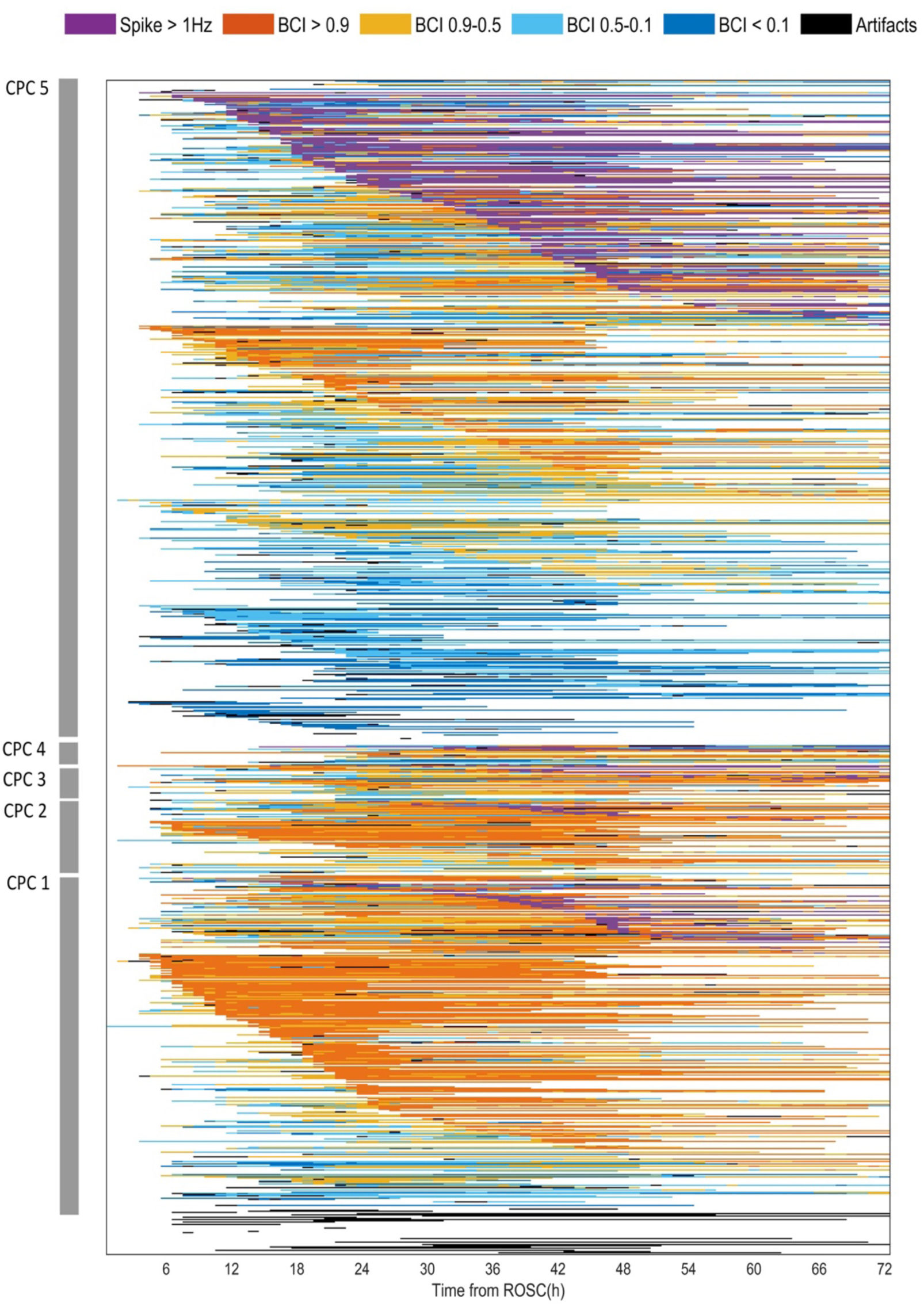
Longitudinal evolution of quantitative EEG categories for individual patients. Patients are ranked based on CPC scores (highest to lowest). First, patients are ranked based on the presence of epileptiform activity (“spike”; purple) and the time of start of epileptiform activity. Patients without epileptiform activity are ranked based on the time of start of a continuous background (orange). Patients without epileptiform activity are ranked based on the presence of a discontinuous, burst suppression, or suppressed background. Patients who predominantly had artifactual EEG data are listed last. CPC: Cerebral Performance Category.

**Table 2.** Number of Patients Per Category Stratified by Neurological Outcome.

|  | **CPC 1** | **CPC 2** | **CPC 3** | **CPC 4** | **CPC5** |
| --- | --- | --- | --- | --- | --- |
| Epileptiform activity >1 Hz | 52 | 17 | 13 | 5 | 171 |
| BCI > 90 % | 212 | 51 | 23 | 9 | 205 |
| BCI 90-50 % | 179 | 47 | 25 | 12 | 307 |
| BCI 50-10 % | 111 | 39 | 21 | 9 | 348 |
| BCI < 10 % | 44 | 13 | 8 | 8 | 241 |

CPC: Cerebral Performance Category
BCI: Background continuity index

## Discussion

The I-CARE consortium database is a large collection of real-world clinical and continuous EEG data from critically ill patients with coma after cardiac arrest from seven institutions in the U.S. and Europe. This database provides a rich ground for advancing critical care neurophysiology research in several ways. First, this disease-specific dataset has a wide range of abnormal EEG patterns in the ictal-interictal continuum and seizures as well as background changes from suppressed to burst suppression and continuous activity. These are of great interest for decision-making about seizure and sedation treatment as well as prognostication based on international specialty guidelines. The size of this dataset allows for more precise estimation of the incidence of epileptiform activity as well as uncertainty in prognostic performance of specific patterns. The EEG patterns post-cardiac arrest are dynamic, evolving in different trajectories based on the time of ROSC (12–14, 18). This database enables testing of models evaluating emergence of seizures and other epileptiform and non-epileptiform patterns or neurological function recovery that leverage these complex time-series’ dynamics. Continuous multi-channel EEG monitoring is prone to artifact and signal quality deterioration given the prolonged duration of recording and context of care in an intensive care unit, therefore this database provides a wealth of different artifact types, which can be used for pursuits focused on artifact detection and rejection as well as development of techniques to enable analyses that are robust to artifact. Finally, the diversity of hospital settings in which these this data were generated provide an opportunity to test algorithm performance across institutions and geographical regions. Advances in algorithm development using traditional signal processing as well as artificial intelligence methods rely on the availability of large and diverse datasets that have been harmonized and organized for research (22). The I-CARE consortium’s effort to support open data may accelerate research in cardiac arrest and clinical neurophysiology as well as other types of acute brain injury.

The illustrative results of epileptiform activity and continuity evolution described here reinforce the temporal dynamics and diversity in EEG patterns seen post-cardiac arrest (14). It also emphasizes the high incidence of epileptiform activity as well as the time-dependence of burst suppression and discontinuity across outcomes (23). These data emphasize previous literature showing that early EEG changes are associated with neurological recovery potential and that improvement in EEG background can be seen in patients with good and poor outcomes, but that the timing of this recovery is prognostic (8, 20). Importantly, caution on interpretation of automated analysis of EEG and use of machine learning and deep learning approaches on continuous EEG data generated in the intensive care units is needed given the high burden of artifactual data seen in these recordings (24).

This database has limitations. Patient care of comatose cardiac arrest patients is complex, and several types of information regarding the clinical context of EEG recording are not available. Temperature modulation and sedative and vasopressor use may influence EEG signals (25, 26). Coexisting comorbidities may contribute to encephalopathy and fluctuating changes on EEG data (27). Variability in clinical practice is common in care of patients with cardiac arrest, which can be present at the provider and institution level (28). Timing of initiation and termination of EEG are highly variable and data availability may be confounded by clinical practice and patient specific factors that are unmeasured. Specifically, bias related to premature predictions of likelihood of neurological recovery and withdrawal of life-sustaining therapies may lead to shortening of EEG recordings as well as affect treatment intensity, which may affect EEG signals (e.g., seizure treatment or presence of burst suppression) (9, 28). Individual level assignment of neurological prognosis and results of other multimodal prognostic testing is not available, therefore determination of the likelihood of bias related to self-fulfilling prophecies from withdrawal of life-sustaining therapies is not possible, particularly because clinicians caring for these patients were not blinded to EEG test results. The EEG recordings included in this study were obtained during routine clinical care and converted using standard clinical software at individual sites without a centralized method, therefore gaps in data due to patient disconnection for transport for procedures and testing as well as data quality issues related to data conversion may be present. While this large dataset has a diverse collection of EEG patterns that has supported previous studies demonstrating that outcome prediction with high specificity can be achieved for a large proportion of patients, there remains opportunity for model performance improvement for a significant fraction of patients. This is particularly relevant for less prevalent subtypes of EEG patterns in patients who recover from coma. The variability in EEG duration and potential bias associated with file duration and withdrawal of life-sustaining therapies may limit deployment of prediction models for time-series using this database. Prospective determination of neurological outcome and use of other patient-centered outcome metrics such as quality of life and cognitive measures was not possible for all participating centers, limiting its application for more nuanced assessment of neurological recovery in this patient population. Near all deceased patients during their initial hospital stay and most patients who recovered did so prior to discharge, therefore the impact of lacking prospective long-term outcome assessment is mitigated, with only 2% of patients required retrospective long-term outcome determination. Informed consent for data sharing was waived, but risk for patient reidentification is low based on the standardized deidentification procedures pursued that included date shifting, grouping of ages above 89, and removal of additional patient identifiers from file headers.

## Conclusion

The I-CARE database provides a large and diverse collection of clinical data and continuous EEG recordings that may enable advancements in cardiac arrest clinical research and algorithm development using EEG more broadly. The scale of this dataset allows for corroboration and reevaluation of current guideline recommendations using EEG information, highlighting uncommon clinical scenarios that have less typical trajectories (e.g., burst suppression and epileptiform activity in patients who have good neurological recovery). This database curation effort may support the development of new standards and best practices to enable multimodal signals data sharing and open access for EEG data that is geared for deployment of traditional signal processing and artificial intelligence methods in the intensive care unit environment.

## Data Availability

All data produced are available online at bdsp.io.

https://bdsp.io/

---

**Figure S1:**
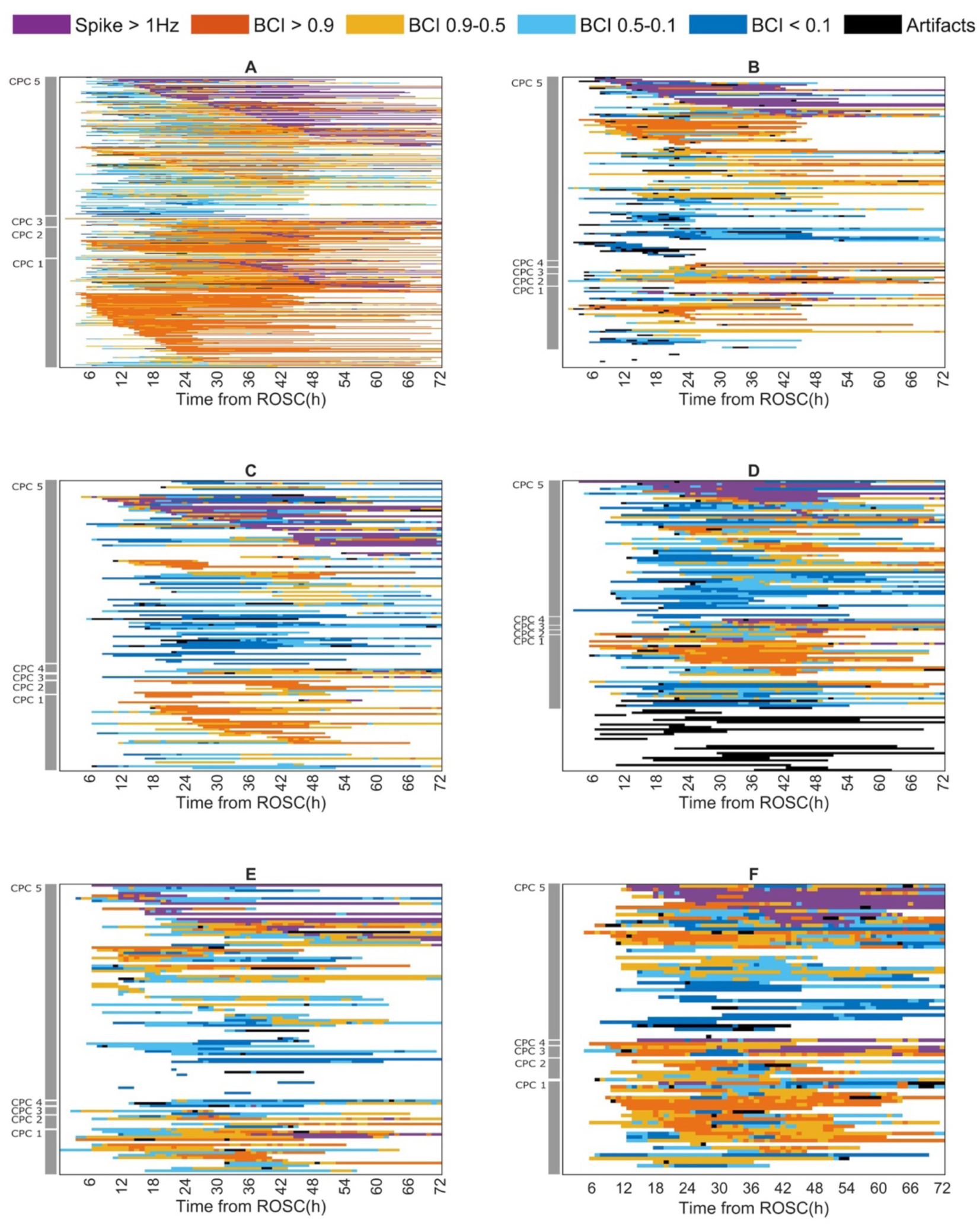
Longitudinal evolution of quantitative EEG categories for individual patients stratified by hospital (A-F). Patients are ranked based on CPC scores (highest to lowest). First, patients are ranked based on the presence of epileptiform activity (purple) and the time of start of epileptiform activity. Patients without epileptiform activity are ranked based on the time of start of a continuous background (orange). Patients without epileptiform activity are ranked based on the presence of a discontinuous, burst suppression, or suppressed background. Patients who predominantly had artifactual EEG data are listed last. CPC: Cerebral Performance Category.

